# Estimating the effects of exercise-based cardiac rehabilitation on health-related quality of life in patients with myocardial infarction in Sweden: A registry-based study using non-experimental methods

**DOI:** 10.1101/2025.01.12.25320433

**Authors:** Anja N. Stanke, Margrét Leósdóttir, Maria Bäck, Björn Ekman

**Author notes:** ANS led the data analysis and the drafting of the manuscript. BE provided analytical support. ML and MB provided support to the conceptualization of the study. All authors contributed to the writing of the manuscript. Corresponding author: Björn Ekman.

## Abstract

Participation in exercise-based cardiac rehabilitation (EBCR) improves long-term prognosis for patients following myocardial infarction (MI). Evaluating the effects of EBCR is challenging due to the voluntary nature of participation in the program. Using register data on 18,978 patients hospitalized for MI in 2011 to 2013 in Sweden, a non-experimental study design is employed to evaluate the effects of EBCR on the health-related quality of life (HRQoL). Conditional independence from non-random treatment assignment is considered through set of potential-outcome (PO) models to estimate the average treatment effect (ATE) and the ATE on the treated (ATET) of EBCR on HRQoL using the EQ-5D-3L instrument. Changes in the EuroQoL Visual Analogue Scale (EQ-VAS; range 0 to 100) and the EQ5D-Index (range -0.594 to 1) between 6-10 weeks and one-year post-MI are compared between two groups of participants in EBCR (complete participants [≥3 months] and non-complete participants [<3 months]) and non-participants (control group). In total, 43% of patients participated in EBCR to some extent (12% complete, 31% non-complete). Participants showed greater improvement on the EQ-VAS scale compared with non-participants. Patients who completed an EBCR program had a larger improvement (ATE of between 1.851-1.917 points; p < 0.001) compared with participants who did not complete the full program (0.974-1.017 points; p < 0.01). Full participation in the program increased the EQ5D-Index of between 0.014 and 0.017 points (p < 0.05), while no statistically significant effect was observed among non-completers. The effect of EBCR on HRQoL was more pronounced among younger patients and those reporting dyspnea and angina than older and non-symptomatic patients. Participating in EBCR may improve the health-related quality of life of patients post-MI. The clinical relevance of the observed impact of EBCR on HRQoL warrants further investigation. The effects of EBCR participation can be readily estimated using broadly applicable non-experimental approaches and commonly available register data.

## 1 Introduction

Individuals who suffer a myocardial infarction (MI) face an elevated risk of repeated events and death [1] as well as significant impairments in health-related quality of life (HRQoL) [2–4]. HRQoL is a multifaceted concept that assesses a person’s overall functioning and well-being across physical, mental, and social dimensions of health [5]. Losses in HRQoL have repeatedly been reported among patients with MI [2–4]. Furthermore, low HRQoL post-MI is associated with an increased risk of repeated hospital admissions [6], major cardiovascular events and death [7–9]. Consequently, secondary preventive measures aiming to prevent recurrent cardiac events and promote HRQoL among survivors of MI are critical to the effective management and care of this patient group.

Guidelines from the European Society of Cardiology for patients with acute coronary syndromes [10] recommend the provision of cardiac rehabilitation (CR) in general, and exercise-based CR (EBCR) in particular, due to its well-established positive effects on cardiovascular risk factor management [11] and prognosis [12]. Although implementation of EBCR programs varies across different settings, such programs generally involve medically supervised physical exercise that combines aerobic and resistance training over a period of at least three months. In Sweden, EBCR participation is routinely offered to post-MI patients [13]. In particular, 24 EBCR sessions for 3 months, along with a pre-exercise and a post-exercise assessment, is recommended.

While extensive research demonstrates the positive effects of EBCR on objectively measured risk factors, previous systematic reviews evaluating the impact of EBCR on HRQoL in patients with coronary artery disease have reported inconsistent results [12, 14–16]. This may stem from differences across studies regarding study populations, length of follow-up, HRQoL measurement tools, content, and dose of EBCR treatment. Although randomized controlled trials (RCTs) are often considered the gold standard in the evaluation of treatment effects [17], there are also methodological constraints to RCTs, including the potential for selection-, performance-, and attrition bias [15, 16]. Moreover, the reliance on small samples in RCTs raises concerns about external validity and generalizability.

Observational studies with representative samples can be beneficial in this context, in particular when applying methods that go beyond mere establishment of an association [18, 19]. To date, only a few well-designed observational studies [20–22] have examined EBCR participation and HRQoL outcome measures. The findings of these studies suggest mixed effects of EBCR on participants’ quality of life. However, none of the studies have solely assessed supervised exercise, but rather considered the impact of a combination of components, including supervised exercise and other treatments, such as dietary and smoking cessation counselling and stress management. Importantly, no study on this issue has applied analytical methods that address the underlying assumption in treatment effect studies of conditional independence between the outcome and treatment assignment. To address this gap in the evidence base, we aimed to evaluate the effect of EBCR on self-reported HRQoL among patients with MI using methods that allow identification of a treatment effect based on observable factors that affect both treatment assignment and outcomes. Specifically, we estimated the average treatment effects (ATE) of EBCR participation by fitting a set of potential-outcome (PO) models to account for self-selection of patients into treatment status. In addition, and building on previous research suggesting that exercise dosage may influence the degree to which physical exercise improves HRQoL [23], we also aimed to investigate whether a dose-response relationship exists between EBCR and HRQoL.

## 2 Materials and methods

### 2.1 Study design and sample

We employed a retrospective cohort study design based on data from the Swedish Web-system for Enhancement and Development of Evidence-based care in Heart disease Evaluated According to Recommended Therapies (SWEDEHEART) [24]. The SWEDEHEART registry collects data on a broad range of patient and treatment characteristics, including demographic information, cardiovascular risk factor management, health behavior, recurrent cardiac events, EBCR participation, and HRQoL. In 2013, 95% of all Swedish CR centers reported to SWEDEHEART, covering 80% of all patients under the age of 75 who had been hospitalized for MI and were alive one year after the acute event [25]. To track the entire patient pathway from acute hospital care to outpatient rehabilitation, SWEDEHEART data is collected on three occasions: at hospitalization (baseline) and at two follow-up visits: 6-10 weeks after MI and 12-14 months after MI. The sample of patients with MI used in this study was drawn from the registered MI cases in SWEDEHEART between 2011-2013 (made available for research purposes on 15/01/2024). In total, 18,978 MI-patients were included, encompassing all patients who had been diagnosed with MI within the three-year period, who attended both follow-up visits, and had non-missing data on EBCR participation.

### 2.2 Study variables and measures

In SWEDEHEART, self-reported HRQoL is measured with the standardized EuroQol-Five-Dimensions-Three-Level Questionnaire (EQ-5D-3L) [26]. The EQ-5D-3L is validated for use among patients with MI [27] and comprises two parts: a descriptive system (EQ5D-Index) and a visual analog scale (EQ-VAS). The EQ5D-Index is a weighted summary score that reflects an individual’s health status across five dimensions (mobility, self-care, usual activities, pain/discomfort, anxiety/depression) while taking into account the health norms of the underlying population. The EQ5D-Index values are benchmarked against the norms of the general population, which in the Swedish case is the UK population norm [28]. The score ranges from negative values (representing a state considered worse than death) to 1 (representing full health).

The EQ-VAS captures an individual’s perception of health status, with participants being asked to rate their current health state on a scale ranging from 0 (“Worst health you can imagine”) to 100 (“Best health you can imagine”) [26]. To evaluate the impact of EBCR on HRQoL, we assessed the change in EQ-VAS score (ΔEQ-VAS) and the change in EQ5D-Index score (ΔEQ5D-Index) between the first and the second follow-up visits.

*Participation in EBCR* until the second follow-up visit served as the multivalued treatment variable. Differentiating between no participation (non-participants), participation for less than the recommended three months (non-completers), and participation for at least three months (completers) allowed for the investigation of a potential dose-response relationship between EBCR and HRQoL.

To model the relationship between EBCR and HRQoL, we utilized information on a range of observable factors known to affect the quality of life of patients with MI from the SWEDEHEART registry, including the age and sex of the patients, employment status, smoking, diabetes, previous MI and/or stroke, dyspnea and/or angina symptoms, and physical activity above average. In addition, an estimate of the distance from the patient’s home to the clinic was linked to the current dataset [11]. This measure was hypothesized to affect participation in the EBCR program [29] but not to affect the outcome. As such, it serves as an instrumental variable in the models enhancing the strength of the analysis.

To avoid underpowered categories, we dichotomized the categorical variables *employment status* and *smoking,* differentiating between employed/non-employed, and smoker/non-smoker. In SWEDEHEART, angina is rated based on the Canadian Cardiovascular Society (CCS) grading of angina pectoris [30], while dyspnea is classified based on the New York Heart Association (NYHA) Functional Classification system [31]. Angina and dyspnea were considered present when patients reported symptoms rated as class II-IV on the CCS and NYHA scales, respectively. The variables *dyspnea* and *angina* were dichotomized and combined into the binary variable *dyspnea and/or angina symptoms* to capture a more comprehensive measure of potential symptoms of a similar nature. The binary variables *previous MI* and *previous stroke* were combined into the variable *previous MI and/or stroke*. Finally, we created the variable *physical activity above average* to indicate whether a patient’s normal physical activity level (measured by the self-reported number of physical activity days per week) was above or below the average observed in the sample.

The selection of the covariates was informed by insights from prior studies on the relationship between EBCR and HRQoL [7, 29, 32, 33]. Data on the covariates *dyspnea and/or angina symptoms* and *number of physical activity days per week* were collected during the first follow-up visit, while information on all other covariates was gathered at the time of MI. An overview of all variables used in the study is given in Table S1.

### 2.3 Statistical methods

To address the possibility of unobserved confounding, we employed potential outcomes models to estimate the average treatment effects (ATE) and the average treatment effects on the treated (ATET) [34]. Specifically, to obtain consistent estimates of the effects of EBCR on HRQoL we used the following potential outcomes treatment effect estimators: regression adjustment (RA), inverse probability weighting (IPW), inverse probability weighting with regression adjustment (IPWRA), and augmented inverse probability weighting (AIPW) [35]. To account for self-selection into treatment groups these approaches utilize observable information to create comparability between the treatment group(s) and an untreated control group [36]. While RA estimators model the outcome and IPW estimators the treatment assignment process, the IPWRA and AIPW predictors embody a doubly-robust property by predicting both the outcome and the treatment assignment process [35]. Doubly-robust estimators come with the advantage that only one model needs to be correctly specified for consistent estimation [35].

Our identification approach involved a series of steps, including: i) obtaining baseline differences between the EBCR groups and the control group using independent samples t-tests and chi-square tests of independence, as appropriate; ii) comparing the observed mean changes in EQ-VAS and EQ5D-Index between treatment groups by 1-year post-MI using independent samples t-tests; iii) controlling for the effect of other variables that may distort the relationship between EBCR and HRQoL; iv) exploring whether the effect of EBCR on HRQoL was strengthened or weakened depending on a patient’s sex, age, dyspnea and/or angina symptoms, and overall physical activity level by conducting interaction analyses; and v) performing sensitivity checks to test the robustness of our findings.

In step iii, we addressed the potential for endogeneity bias by a) including age, sex, employment status, dyspnea and/or angina symptoms, diabetes, smoking, and number of physical activity days per week as potentially confounding factors in multiple linear regression models (hereinafter referred to as “base models”, see Table S2 and S3) and b) employing the previously described potential outcomes treatment effect estimators: RA, IPW, IPWRA, and AIPW. To model the outcome, we utilized the covariates age, sex, employment status, dyspnea and/or angina symptoms, diabetes, smoking, and number of physical activity days per week. The same covariates along with the variable distance to clinic were used to predict treatment allocation. The variable smoking was only included in the EQ-VAS models, as Wald test results indicated that including smoking in the EQ5D-Index models would increase the error term without substantially improving the prediction. All data management and analyses were done using Stata 17.

### 2.4 Ethical considerations

The study complied with the Declaration of Helsinki and was approved by The Regional Ethical Review Board at Lund University, Sweden (Registration numbers 2014/6 and 2014/387). The need for informed participant consent was waived by the ethics review board as all data were anonymized and the researchers did not have access to information that could identify individual participants during or after the implementation of the study.

## 3 Results

### 3.1 Descriptive statistics

The total sample included almost 19,000 patients, around 12 percent of whom participated fully in the EBCR program (completers) and 31 percent participated albeit <3 months (non-completers). Approximately one-third of the sample was female, and the sample’s average age was 63 years (s.d. 8.66 years), 42 percent were employed, and 29 percent were current smokers at the time of hospitalization. With respect to comorbidities, around one-fifth of the sample had suffered from previous MI and/or stroke, and 18 percent had diabetes at the time of hospitalization. On average, patients reported being physically active on four days a week at the time of the first follow-up visit. A detailed description of the study sample can be found in Table 1.

**Table 1.** Descriptive statistics at baseline by participation status.

|  | **Total**<br>**18,978** | **No participation**<br>**N = 10,761** | **Participation**<br>**< 3 months**<br>**N=5,841** | **Participation**<br>**≥ 3 months**<br>**N=2,376** |
| --- | --- | --- | --- | --- |
| Age, years |  |  |  |  |
| Mean | 62.78 | 63.22 | 61.83 | 63.17 |
| SD | 8.66 | 8.68 | 8.69 | 8.33 |
| Male |  |  |  |  |
| N | 14,180 | 8,085 | 4,338 | 1,757 |
| % | 74.72 | 75.13 | 74.27 | 73.95 |
| Employed at baseline |  |  |  |  |
| N | 7,598 | 3,994 | 2,650 | 954 |
| % | 42.31 | 39.27 | 47.76 | 42.63 |
| Previous MI and/or stroke |  |  |  |  |
| N | 3,765 | 2,506 | 889 | 370 |
| % | 19.93 | 23.40 | 15.28 | 15.64 |
| Dyspnea and/or angina symptoms at 1<sup>st</sup> follow-up |  |  |  |  |
| N | 2,032 | 1,209 | 568 | 255 |
| % | 12.20 | 13.17 | 10.61 | 11.96 |
| Diabetes at baseline |  |  |  |  |
| N | 3,413 | 2,171 | 871 | 371 |
| % | 18.02 | 20.21 | 14.96 | 15.65 |
| Smoker at baseline |  |  |  |  |
| N | 5,405 | 3,390 | 1,510 | 505 |
| % | 29.10 | 32.16 | 26.42 | 21.79 |
| Number of physical active days per week at 1<sup>st</sup> follow-up |  |  |  |  |
| Mean | 4.05 | 3.73 | 4.45 | 4.47 |
| SD | 2.65 | 2.77 | 2.44 | 2.46 |
| Distance to clinic, km |  |  |  |  |
| Mean | 22.04 | 23.23 | 20.25 | 20.98 |
| SD | 10.37 | 10.86 | 9.27 | 9.88 |

### 3.2 Baseline differences between treatment groups

Table 1 shows the baseline differences between the two groups of EBCR participants and the control group for all covariates. While some differences between non-completers (EBCR <3 months) and completers (EBCR ≥3 months) were evident, more notable differences were observed between those attending EBCR and those not attending EBCR. Patients not attending EBCR were more likely to be smokers, to be diagnosed with diabetes, to have experienced previous MI and/or stroke, and to report dyspnea and/or angina symptoms compared with patients attending EBCR. Also, non-participants were more often male, living further away from the clinic, and reported a slightly more physically active lifestyle than participants. All observed intergroup differences, except for gender distribution, were statistically significant (Table S4 and S5).

#### 3.2.1 Differences in HRQoL between and within treatment groups

The EQ5D-Index values of participants (completers and non-completers combined) and non-participants at the first and second follow-up visits are presented in Figure 1.

**Figure 1.**
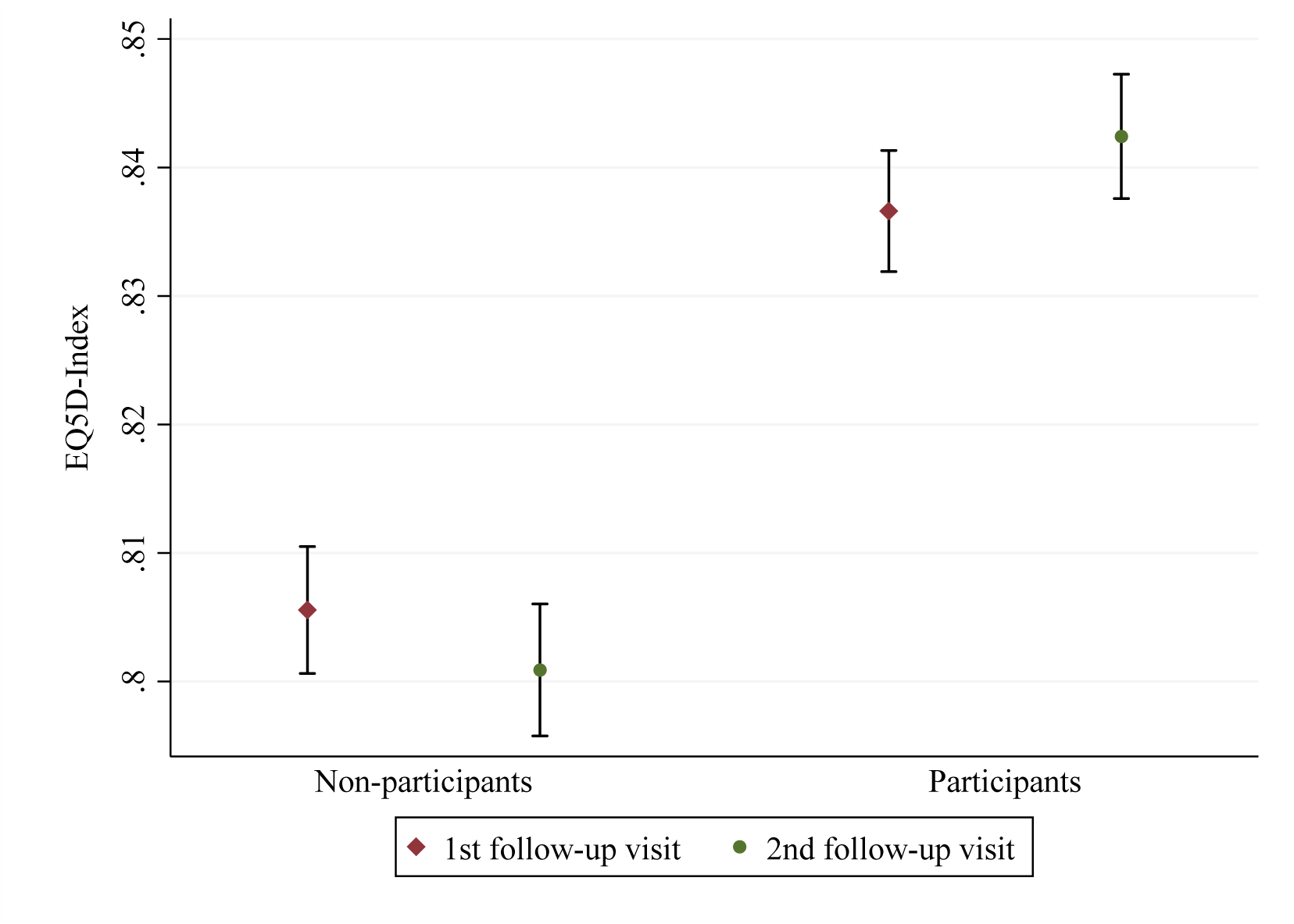
EQ5D-Index scores with 95% Confidence Intervals at the 1^st^ and 2^nd^ follow-up visits by treatment group.

While among participants the EQ5D-Index score increased slightly between the follow-up visits (p=0.022), no such change in the EQ5D-Index score was observed for non-participants (p=0.061). Employing independent samples t-tests, the observed difference between EBCR participants and non-participants in ΔEQ5D-Index was statistically significant (p=0.003). Intergroup differences for EQ-VAS are shown in Figure 2. Over the one-year follow-up period non-participants reported a mean increase of 1.5 points (p=0.0000) while EBCR participants reported a mean increase of 2.9 points on the EQ-VAS scale (p =0.0000).

**Figure 2.**
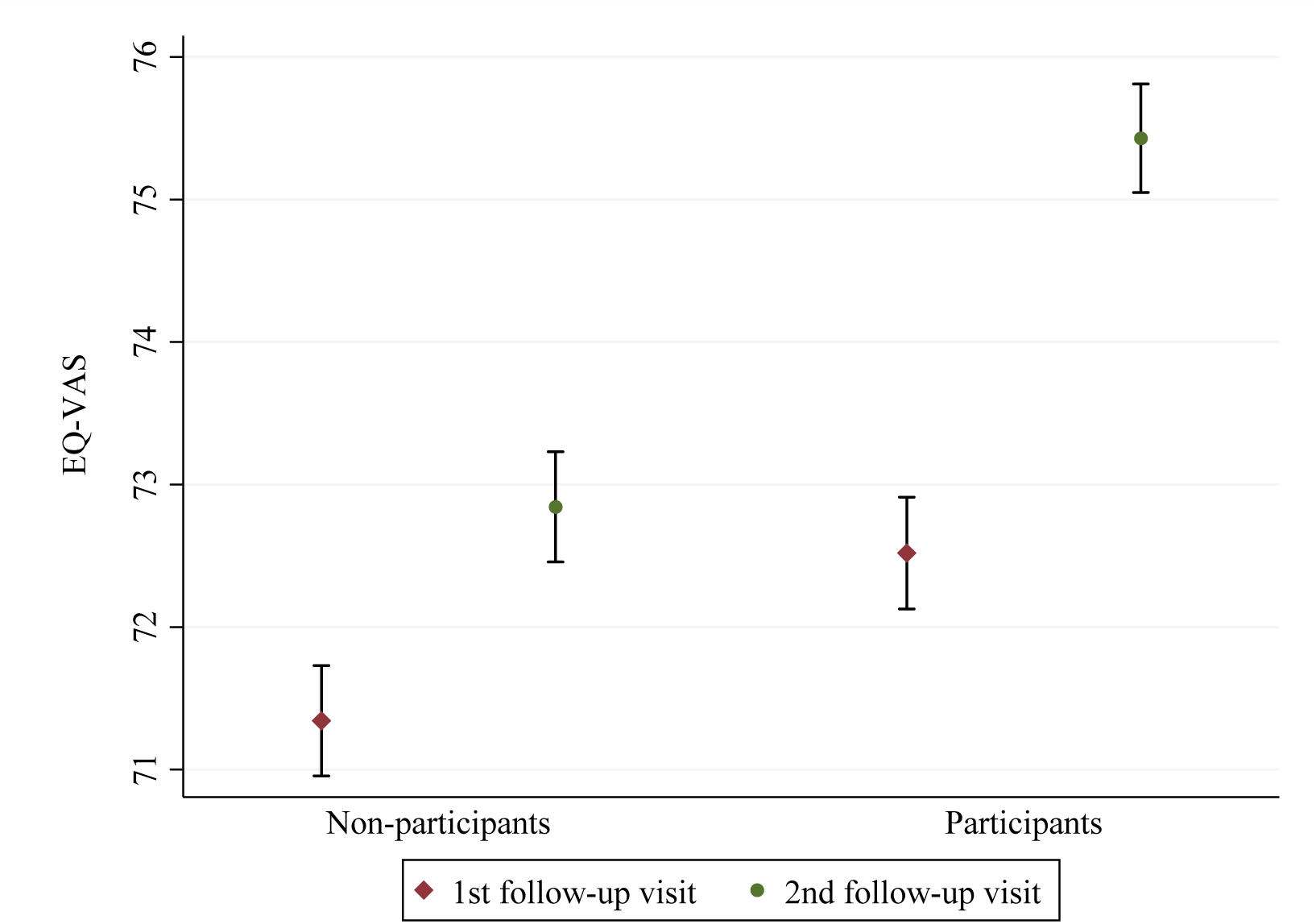
EQ-VAS scores with 95% Confidence Intervals at the 1^st^ and 2^nd^ follow-up visits by treatment group.

In addition to intergroup differences, we also found differences in HRQoL status within the EBCR treatment group. Specifically, we found that EBCR participants suffering from dyspnea and/or angina symptoms had lower HRQoL scores at the first follow-up visit (EQ-VAS: 61.6±18.3; EQ5D-Index: 0.73±0.24) compared to non-symptomatic EBCR participants (EQ-VAS: 73.9 ±16.7; EQ5D-Index: 0.85±0.2) (p=0.0000). Similarly, HRQoL values at the first follow-up visit were found to be lower among younger EBCR participants than among older participants (Table S6).

### 3.3 Potential-outcomes models

Table 5 shows the estimated treatment effects of EBCR on ΔEQ-VAS from fitting the RA, IPW, IPWRA, and AIPW models; ATE upper part and ATET lower part. Compared to non-participation, participation in EBCR was associated with an increase in the absolute EQ-VAS score by between 0.974 and 1.017 points among non-completers (EBCR < 3 months) and by between 1.851 and 1.917 points among completers (EBCR ≥ 3 months), after adjusting for observable factors. The ATET of EBCR on EQ-VAS took on slightly smaller values and ranged from 0.797 to 0.887 points among non-completers and from 1.733 to 1.857 points among completers. All effect measures were statistically significant (Table 5) and aligned with the findings of the base models (Tables S2 and S3), suggesting a significant albeit small effect of EBCR on ΔEQ-VAS.

**Table 5.** Estimated changes in EQ-VAS scores at one-year post-MI (ATE and ATET).

|  | RA | IPW | IPWRA | AIPW |
| --- | --- | --- | --- | --- |
| **ATE** |  |  |  |  |
| EBCR < 3 months | 0.974**<br>[0.331; 1.616] | 0.994**<br>[0.328; 1.660] | 1.017**<br>[0.350; 1.683] | 1.014**<br>[0.347; 1.680] |
| EBCR ≥ 3 months | 1.917***<br>[1.020; 2.815] | 1.851***<br>[0.921; 2.782] | 1.852***<br>[0.923; 2.781] | 1.851***<br>[0.922; 2.781] |
| *N* | 15,422 | 15,150 | 15,150 | 15,150 |
| **ATET** |  |  |  |  |
| EBCR < 3 months | 0.887**<br>[0.257; 1.518] | 0.809*<br>[0.154; 1.464] | 0.797*<br>[0.142; 1.451] | - |
| EBCR ≥ 3 months | 1.857***<br>[0.985; 2.729] | 1.743***<br>[0.850; 2.637] | 1.733***<br>[0.841; 2.626] | - |
| *N* | 15,422 | 15,150 | 15,150 | - |

Notes: 95% confidence intervals in brackets (based on robust standard errors). \* $p < 0.05$ , \*\* $p < 0.01$ , \*\*\* $p < 0.001$ . Acronyms: RA stands for regression adjustment, IPW stands for inverse probability weights, IPWRA stands for inverse-probability-weighted regression adjustment, AIPW stands for augmented inverse-probability weights.

The corresponding results for the EQ5D-Index measure of HRQoL are presented in Table 6. For those who participated less than the recommended three months in the EBCR program no effect was found compared with those who did not participate at all. In contrast, those who completed the full program did experience a statistically significant effect of the program compared with those who did not participate. Compared to the ATE values, the ATET values were statistically significant for both treatment groups, amounting to 0.013 points among completers and ranging from 0.009 to 0.010 points among non-completers.

**Table 6.** Estimated changes in EQ5D-Index at one-year post-MI (ATE and ATET).

|  | RA | IPW | IPWRA | AIPW |
| --- | --- | --- | --- | --- |
| ATE |  |  |  |  |
| EBCR < 3 months | 0.008<br>[-0.000; 0.016] | 0.008<br>[-0.001; 0.016] | 0.008<br>[-0.000; 0.016] | 0.008<br>[-0.000; 0.016] |
| EBCR ≥ 3 months | 0.014*<br>[0.002; 0.025] | 0.017**<br>[0.005; 0.029] | 0.017**<br>[0.005; 0.029] | 0.017**<br>[0.005; 0.029] |
| *N* | 15,737 | 15,458 | 15,458 | 15,458 |
| ATET |  |  |  |  |
| EBCR < 3 months | 0.009*<br>[0.002; 0.017] | 0.010*<br>[0.002; 0.018] | 0.010*<br>[0.002; 0.018] | - |
| EBCR ≥ 3 months | 0.013*<br>[0.002; 0.024] | 0.013*<br>[0.002; 0.024] | 0.013*<br>[0.002; 0.024] | - |
| *N* | 15,737 | 15,458 | 15,458 | - |

Notes: 95% confidence intervals in brackets (based on robust standard errors). \* $p < 0.05$ , \*\* $p < 0.01$ , \*\*\* $p < 0.001$ . See Table 5 for model acronyms.

To assess the validity of our treatment effect estimations, we tested whether there was an overlap in the covariate distributions between the control group and the treatment groups. Treatment group overlap is a key assumption underlying potential outcome models [34] and was found to be satisfied in the present study (Figures S1 and S2).

#### 3.3.1 Heterogenous and marginal effects

Assuming that the effect of EBCR on HRQoL may not solely depend on the amount of time individuals spend on EBCR participation but also on other patient characteristics, we conducted interaction analyses using the base models described earlier (Table S2-S3). The results suggest that the effect of EBCR on ΔEQ-VAS did not depend on sex, age, and self-reported physical activity while being dependent on the presence of dyspnea and/or angina symptoms (Table S7). Specifically, while all measures were statistically significant, the estimated average change in EQ-VAS score appeared to be notably bigger in EBCR participants with dyspnea and/or angina symptoms (estimated change: 6.20 - 7.32 points) than in EBCR participants without these symptoms (estimated change: 2.02 - 2.94 points; Figure 3).

**Figure 3.**
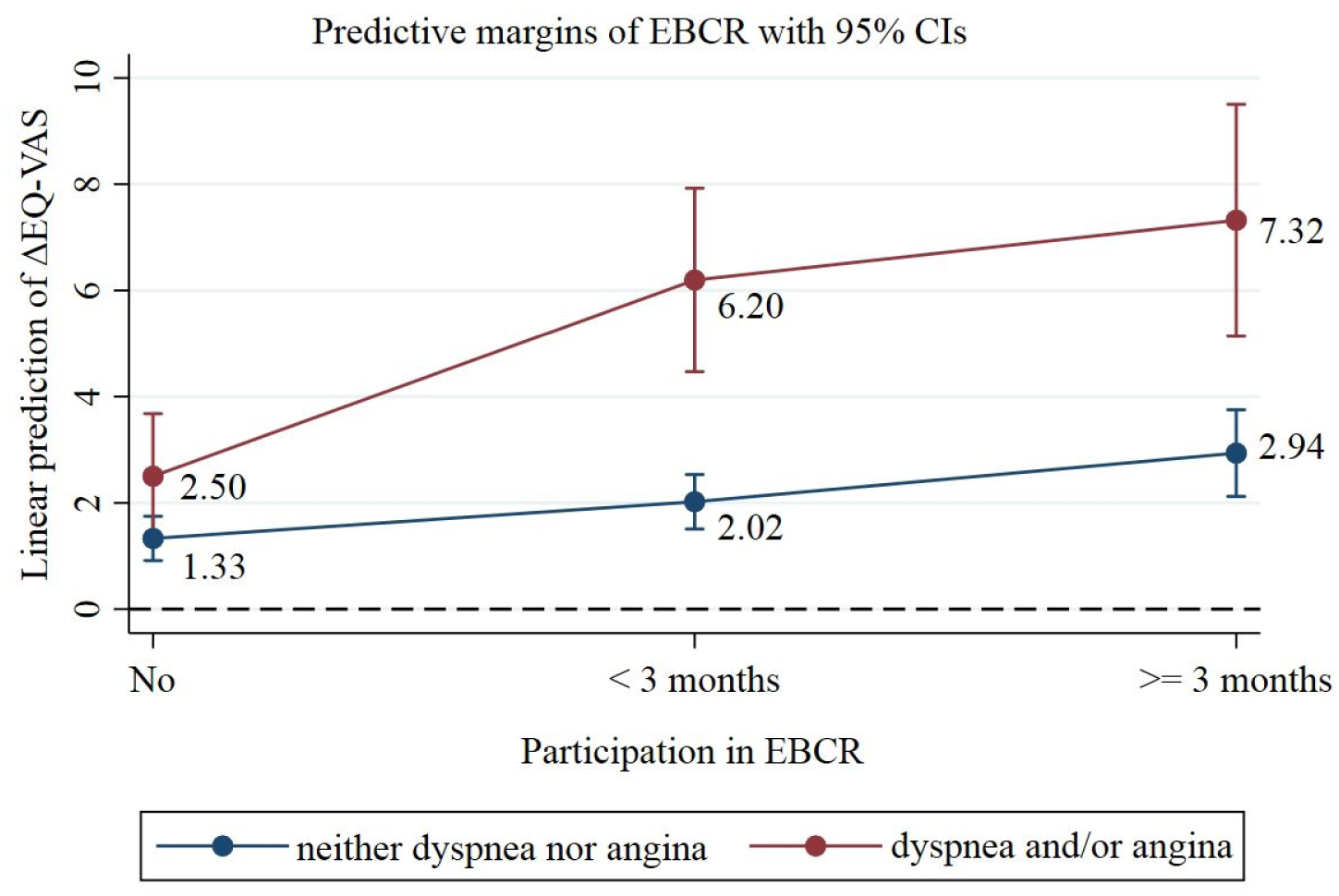
Marginal effects of EBCR on ΔEQ-VAS by symptoms of dyspnea and/or angina. Notes: Angina was considered present when symptoms were rated as class II-IV on the Canadian Cardiovascular Society (CCS) grading of angina pectoris. Dyspnea was considered present when symptoms were rated as class II-IV on the New York Heart (NYHA) Association Functional Classification system.

The effect of EBCR on ΔEQ5D-Index was found to be independent of dyspnea and/or angina symptoms as well as patients’ self-reported physical activity levels (Table S8). In contrast, the improvement in EQ5D-Index over the one-year period appeared to decline with increasing age, eventually losing statistical significance among patients aged 65 years and older (Figure 4).

**Figure 4.**
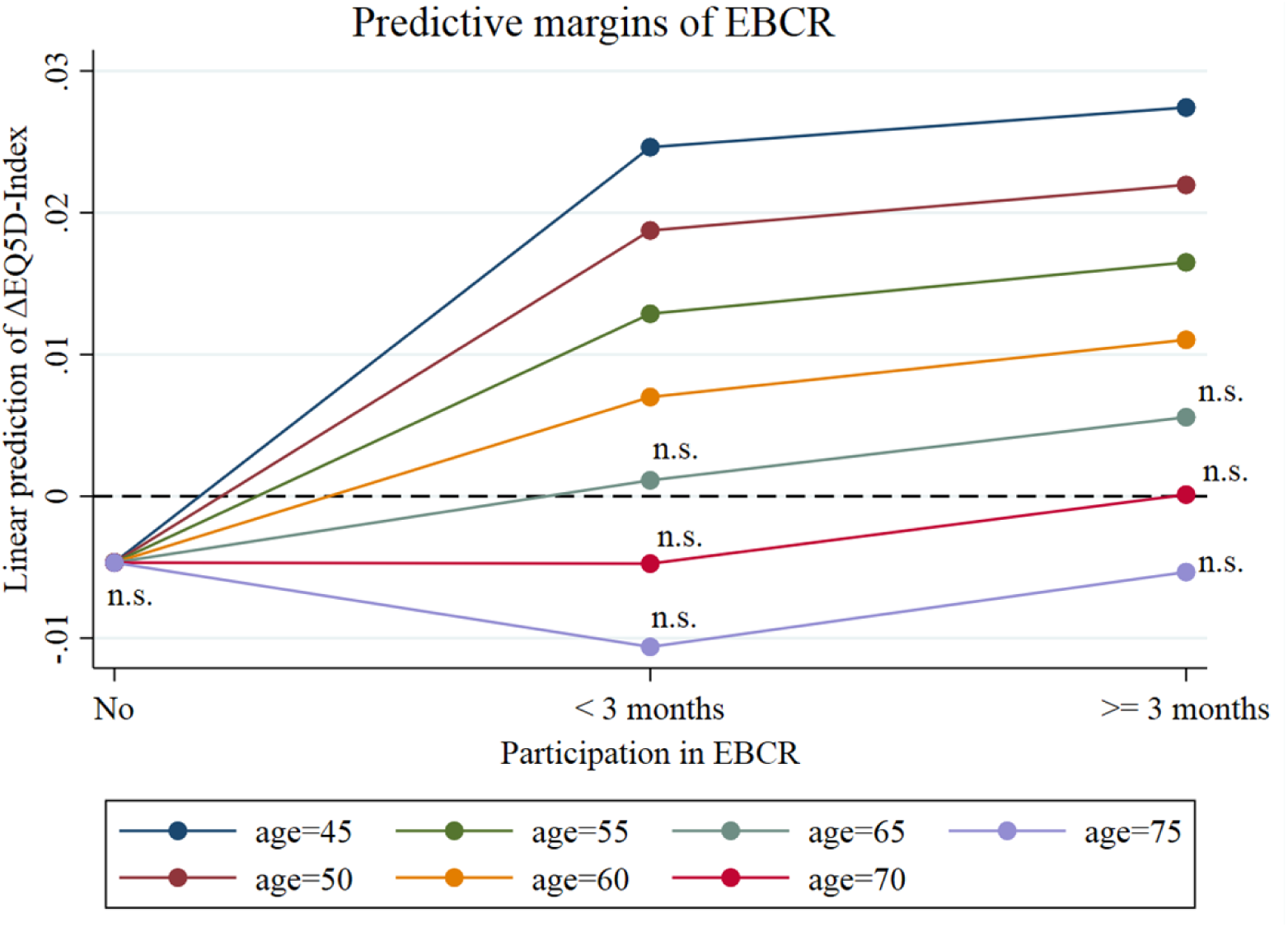
Marginal effects of EBCR on ΔEQ5D-Index by age.

### 3.4 Sensitivity analyses

Adjusting the estimated standard errors for clustering by grouping patients treated within the same clinic into the same cluster, altered some of the reported findings. While the results of the potential outcomes models suggested an effect of EBCR≥3 months on ΔEQ5D-Index (Table 6), neither EBCR<3 months nor EBCR≥3 months were significantly associated with ΔEQ5D-Index after employing clustered standard errors (Table S9-S10). In contrast, the findings on ΔEQ-VAS remained unaffected by clustering (not shown). Excluding patients over 74 years of age from the analyses did not alter the reported findings in any material way (not shown). To investigate the robustness of our findings we also refitted the models while removing ΔEQ-VAS values over ±90 and ΔEQ5D-Index values over ±0.9. This did not change the results in any significant way (not shown).

## 4 Discussion

In this study we examined the effect of EBCR on HRQoL using nationwide registry data on Swedish patients with MI. For this purpose, we gradually employed more evolved regression modelling techniques to address the non-randomized selection into treatment groups. In doing so, this study demonstrates the ability to evaluate the effects of EBCR participation on relevant outcomes in a non-experimental setting by applying a non-experimental approach.

The results of our treatment effect analyses suggest a positive effect of EBCR on the HRQoL of patients with MI. Specifically, we found that EBCR participants experience a statistically significantly greater improvement in EQ-VAS within one-year post-MI than their non-participating counterparts. In finding an effect of EBCR on EQ-VAS, our findings are in line with a recent RCT by Campo et al. [37]. The authors of this study assessed the effect of a combined center-based and home-based exercise program among elderly patients with acute coronary syndrome in Italy and found a statistically significant effect on EQ-VAS, in favor of EBCR participants. In contrast, no effect of EBCR on EQ-VAS was observed in two earlier Swedish RCTs [38, 39], which assessed the effect of supervised exercise on a small group of elderly patients hospitalized for acute coronary events. Using almost the same sample the authors found no significant effect of EBCR on EQ-VAS, neither at one-year [38] nor at 3-6 years follow-up [39]. The contradictory results may arise from differences in sample selection, study design and statistical methods between the reported studies. For example, while our study benefitted from the high coverage of the SWEDEHEART registry, the two earlier Swedish RCTs included mainly elderly male patients, who were hospitalized for MI but also for other coronary events, diminishing the comparability to our findings.

Regarding EQ5D-Index, we found that EBCR completers experienced a greater increase in their EQ5D-Index score over the one-year follow-up period than their non-participating counterparts. In contrast, no such effect was observed for non-completers, suggesting that the effect of EBCR on HRQoL may be dose dependent. However, after adjusting the estimated standard errors for clustering at the clinic level, the effect previously observed for EBCR completion was no longer statistically significant. This points to the possibility that variations in EBCR treatment across clinics may partly explain the relationship between EBCR and HRQoL. Indeed, variations in the availability, quality, content, and duration of EBCR programs in Sweden have been previously reported [40]. Therefore, the possibility of a cluster effect due to intraclass correlations among patients treated within the same hospital appears feasible. However, despite finding no overall effect of EBCR on EQ5D-Index after accounting for differences between CR clinics, there may still be an effect of EBCR on EQ5D-Index within certain subgroups. Specifically, our heterogeneous effect analyses revealed statistically significant marginal effects for EQ5D-Index among younger EBCR participants. This finding suggests that younger patients may derive greater HRQoL-related benefits from engaging in EBCR than older patients.

Inconsistent results regarding the effect of EBCR on EQ5D-Index have also been reported in earlier studies. Namely, while the previously described Swedish RCTs [38, 39] found no effect on EQ5D-Index at one-year [38] or 3-6 years after the coronary event [39], the RCT by Campo et al. [37], found significantly greater improvements in all EQ-5D dimensions except for self-care among program attendees. However, Campo et al. did not report an overall EQ5D-Index score and exclusively enrolled older patients with acute coronary syndrome and restricted physical capabilities, limiting the comparability to our study.

In finding an overall effect for EQ-VAS but not for EQ5D-Index, this study aligns with previous systematic reviews [14, 15] reporting contradictory findings on the effect of EBCR on HRQoL, depending on the HRQoL measurement tool used and the HRQoL dimensions explored. For instance, an RCT using visual analogue scales other than EQ-VAS to assess the effect of EBCR on HRQoL in a sample of predominantly male patients with MI found significantly greater improvements in HRQoL among EBCR participants compared to non-participants [41]. Based on the findings of the present study, it therefore cannot be ruled out that the estimated treatment effect of EBCR might have differed if HRQoL measurement tools other than the EQ-5D-3L questionnaire had been used.

The current study suggests a dose-response relationship between EBCR and HRQoL with completers deriving greater benefits from EBCR than non-completers. This aligns with prior research reporting better HRQoL outcomes with increasing levels of activity [23]. Besides treatment dosage, disease severity and patient age seem to play a role in determining the extent to which EBCR impacts HRQoL. Specifically, the results of our heterogenous effect analyses suggest that EBCR participants with dyspnea and/or angina symptoms experience larger improvements in EQ-VAS than participants without these symptoms.

The more pronounced limitation in HRQoL reported at the first follow-up by participants with dyspnea and/or angina symptoms as well as younger participants may be one reason for their higher responsiveness to the EBCR program. On the other hand, the observed variations in effect size between participant subgroups may also result from the EQ-5D-3L questionnaire becoming less sensitive in detecting actual changes in HRQoL as HRQoL values increase [42]. This phenomenon, referred to as “ceiling effect” [43], appears feasible in the current context, given that participants with dyspnea and/or angina symptoms as well as participants of younger age reported lower HRQoL values at the first follow-up visit compared to their symptom-free and older counterparts. In finding no statistically significant marginal effects of EBCR on EQ5D-Index among older participants, our study aligns with previous research by Ståhle et al. [44] and Sandström et al. [38], who found no effect of EBCR on EQ5D-Index among elderly MI patients in Sweden. In contrast Campo et al. [37] reported a significant effect of EBCR on all EQ5D dimensions (except for self-care) in 70+-year-old patients with acute coronary syndrome and limited physical performance. However, considering that their patients had notably lower baseline HRQoL levels compared to the elderly patients in our study, their patients might have been more receptive to the positive effects of EBCR, and ceiling effects may have been less of a concern, possibly explaining the conflicting findings.

Overall, we found a statistically significant yet small effect of EBCR participation on the change in EQ-VAS during follow-up. Whether this effect is clinically meaningful is difficult to answer given that there is no established minimum clinically important difference (MCID) for EQ-VAS in patients with MI. However, given that previous studies on patients with chronic diseases other than cardiovascular conditions have reported larger MCIDs for EQ-VAS, ranging from 6.9 to 10.8 points [45–47], the clinical meaningfulness is to be questioned. In contrast, the effect of EBCR found among participants with dyspnea and/or angina symptoms (marginal means ranging from 6.2 to 7.32 points for EQ-VAS, Figure 3) approached the above mentioned MCIDS, potentially indicating clinical meaningfulness in this patient subgroup. The statistically significant marginal effects of EBCR on EQ5D-Index scores observed among participants aged 60 and younger were also found to be small, again raising concerns about their clinical meaningfulness.

The current study sought to address the challenges arising from the non-randomized allocation of EBCR treatment by utilizing potential outcomes models. Although these models allowed for a more precise treatment effect estimation by controlling for observable factors, the causal connection between EBCR and HRQoL remains unclear, as potential distortions from self-selection bias cannot be completely ruled out. For instance, previous studies found that indicators of socioeconomic status are associated with both EBCR attendance [48] and HRQoL [49]. In the current study, we did not have access to data on socioeconomic status, introducing the possibility of unobserved confounding. Therefore, more advanced statistical techniques accounting for the possibility of unobserved confounding, such as instrumental variable techniques, should be considered for future research.

A limitation of the SWEDEHEART dataset is that information on the covariates *number of physically active days per week*, and *dyspnea and/or angina symptoms*, as well as on EQ-VAS and EQ5D-Index is only collected at the 1^st^ follow-up and 2^nd^ follow-up visit, while no measurements for these variables are performed during hospitalization. Moreover, at the time of the study information on when patients actually started their EBCR program was not collected in SWEDEHEART [13]. In benefiting from the high coverage of SWEDEHEART, this study more closely reflected the actual MI population than previous highly selective RCTs. Nevertheless, it should be noted that our findings may not be generalizable to patients over 74 years of age, due to the age limit for mandatory registration of patients in SWEDEHEART being set at 74 years during the study period [50].

## 5 Conclusions

Participation in EBCR after suffering from an MI may improve patients’ HRQoL, especially in younger patients and those with symptoms of dyspnea and/or angina. By applying rigorous quasi-experimental methods to readily available register data, this study added to the existing evidence of the effect of EBCR on patients’ HRQoL after MI, while aiming to address the methodological challenges arising from self-selection into EBCR. Given the observed variability in the effect of EBCR on HRQoL across patient subgroups, further research into the impact of EBCR after MI is warranted. To further advance our understanding of this topic, future studies should address the methodological shortcomings of previous research, such as limited generalizability and selection bias. Importantly, our study demonstrates the ability to evaluate the effects of a program such as EBCR to which patients self-select using broadly available register data.

## Data Availability

Data cannot be shared publicly because of confidentiality. Data are available from the authors for researchers who meet the criteria for access to the data.

